# Risk factors associated with morbidity and mortality outcomes of COVID-19 patients on the 14^th^ and 28^th^ day of the disease course: a retrospective cohort study in Bangladesh

**DOI:** 10.1101/2020.08.17.20176586

**Authors:** M.Z. Islam, B.K. Riaz, ANMS Islam, F. Khanam, J. Akhter, R. Choudhury, N. Farhana, N.A. Jahan, M.J. Uddin, S.S. Efa

## Abstract

**Summary:** Diverse risk factors intercede the outcomes of COVID-19. We conducted this retrospective cohort study to identify the risk factors associated with morbidity and mortality outcomes with a cohort of 1016 COVID-19 patients diagnosed in May 2020. Data were collected by telephone-interview and reviewing records using a questionnaire and checklist. Morbidity (64.4% Vs. 6.0%) and mortality (2.3% Vs. 2.5%) outcomes varied between the 14^th^ and 28^th^ day. Morbidity risk factors included chronic obstructive pulmonary disease (COPD) (RR=1.19, RR=2.68) both on the 14th and 18^th^ day while elderly (AOR=2.56) and smokeless tobacco (SLT) (AOR=2.17) on the 28^th^ day. Mortality risk factors included elderly (AOR=10.14), COPD (RR=5.93), and SLT (AOR=2.25) on the 14^th^ day, and elderly (AOR=24.37) and COPD (RR=2.72) on the 28^th^ day. The morbidity risk was higher with chronic kidney disease (CKD) (RR=3.33) and chronic liver disease (CLD) (RR=3.99) on the 28^th^ day. The mortality risk was higher with coronary heart disease (RR=4.54) and CLD (RR=9.66) on the 14^th^ while with diabetes mellitus (RR=3.08, RR=2.08), hypertension (RR=3.14, RR=2.30), CKD (RR=8.97, RR=2.71), and malignant diseases (RR=10.29) on both 14^th^ and 28^th^ day. We must espouse program interventions considering the morbidity and mortality risk factors to condense the aggressive outcomes of COVID-19.

## Introduction

A newly emergent coronavirus (SARS-CoV-2) causes coronavirus disease 2019 (COVID-19) was first documented in Wuhan City, Hubei Province, China in December 2019 as an outbreak of pneumonia of mysterious cause [1]. Based on phylogeny, taxonomy, and established practice, on 11 February 2020, the World Health Organization (WHO) named the disease as COVID-19[2]. WHO declared COVID-19 as a global emergency on 30 January 2020 [3] and as pandemic on 11 March 2020 [4]. Globally 213 countries are confronting the grave consequences of the ongoing COVID-19 pandemic. The situation is sprouting rapidly with increasing case counts and deaths worldwide [5]. In this course, Bangladesh is also confronting the tolls of morbidity and mortality posed by this highly infectious disease with community transmission across the country.

The clinical spectrum of COVID-19 appears to be in a wide range, encompassing asymptomatic infection, mild (40%) or moderate (40%) disease, severe disease (15%) that requires oxygen support, and only 5% critical disease [1]. The incubation period of COVID-19 infection is around 5.2 days and the period from the onset of symptoms to death ranges from 6 to 41 days with a median of 14 days [6]. A study conducted in Wuhan of China found that increased age and various comorbidities like hypertension and diabetes mellitus (DM) were associated with the severity of COVID-19. But the study didn’t identify the tobacco use and chronic obstructive pulmonary disease (COPD) as the risk factors for COVID-19 [7].

Another study in China found that nearly half of the patients had comorbidity where hypertension was the most common followed by DM and coronary heart disease (CHD). The study also established the association between increased age and death of the COVID-19 patients[8]. Another study showed that severe patients were older and had comorbidities including hypertension (30.0%), diabetes mellitus (12.1%), and cardiovascular diseases. The median age was 64 years in severe cases and 51.5 years in non-severe cases [7]. The presence of any comorbidity was more common among the severe patients than those having a mild or moderate disease (38.7% vs. 21.0%) with a similar exposure history between the two groups of disease severity [9].

Widespread shreds of evidence have emphasized the harmful impact of tobacco use possibly related to adverse outcomes of COVID-19. One of the leading studies conducted in the Wuhan city of China found higher percentages of current and former tobacco users among the patients that needed ICU support, mechanical ventilation or who had died, and a higher prevalence of smoking among the severe cases [10].

A study conducted on outcomes of the COVID-19 patients found that non-survivors were more often older and men, and they had a higher prevalence of DM, hyperlipidemia, and CHDs. The history of current tobacco use and having COPD was more among the non-survivors [11]. However, the pandemic is still under progression, diverse risk factors influence the outcomes of COVID-19, but relevant data and studies are very scarce in Bangladesh. Therefore, it is irrefutably obligatory to determine the risk factors to avert the aggressive consequences of COVID-19 patients. Based on these realities, in this particular study, we aimed to identify the risk factors associated with morbidity and mortality outcomes of COVID-19 patients.

## Methods

### Study setting, design, and population

This observational retrospective cohort study was conducted on the National Institute of Preventive and Social Medicine (NIPSOM), Dhaka, Bangladesh during the period from March to June 2020. The study enrolled a cohort of laboratory-confirmed 1016 COVID-19 (SARS-CoV-2) patients diagnosed at the central laboratory of NIPSOM, Dhaka. The NIPSOM is the apex public health institute holding the central laboratory designated for COVID-19 diagnosis, approved by the Ministry of Health and Family Warfare, the government of Bangladesh. The participant who had no contact telephone/cell phone number; who didn’t respond to a phone call on three (03) separate occasions within 28 days of diagnosis; who were unwilling; and who had incomplete interview were excluded from the study.

### Cohort

The cohort comprised of all the laboratory-confirmed COVID-19 patients, who were diagnosed at the central laboratory of NIPSOM during the period from 01 to 30 May 2020 by real-time reverse transcriptase-polymerase chain reaction (RT-PCR) assay of nasopharyngeal (NP) / Oropharyngeal (OP) / Nasal swab.

### Exposures

The exposures were the risk factors linked with the morbidity and mortality outcomes of the COVID-19 patients, which included baseline characteristics, comorbidity, and tobacco use.

### Outcomes

Both morbidity (cure/not-cured) and mortality (survivors/non-survivor) outcomes of the COVID-19 were identified and compared between exposed and non-exposed groups on the 14^th^ and 28^th^ day of the disease course.

### Sample size and sampling

Initially, all the 1187 laboratory-confirmed COVID-19 patients diagnosed in May 2020 were selected as the study cohort. Finally, 1016 COVID-19 patients were enrolled as the study samples considering the selection criteria and single-centered cluster sampling technique. All the COVID-19 patients formed the sampling frame and each patient was a sampling unit.

### Data collection and analysis

Data were collected by telephone interview and medical records review using a semi-structured questionnaire and checklist respectively. Each telephone interview session was recorded by a digital recorder to ensure the validity of data. The data collection instruments were pretested on the COVID-19 patients diagnosed in April 2020 and accordingly necessary corrections were performed for finalization. Participation of the COVID-19 patients was voluntary and informed consent was obtained from each participant before data collection. In case of non-survivor, data were collected from the eligible family member of the respective non-survivor. Measures were taken to ensure data quality; inconsistency and irrelevance of data were checked and corrected. Data were analysed using SPSS STATISTICS (Version 25.0, IBM Statistical Product and Service Solutions, Armonk, NY, USA).

### Statistical analysis

The normality of the variables was tested with the Shapiroe Wilk test/Kolmogorov Smirnov tests of Normality. Continuous data were written in the form of mean and standard deviation. Categorical data were reported as counts and percentages. Descriptive statistics estimated mean, standard deviation, and frequency while inferential statistics included chi-square test, logistic regression, Relative Risk (RR), and Attributable Risk (AR). A p-value < 0.05 was considered significant. All the statistical tests were two-sided and were performed at a significance level of α = 0.05.

### Measurement of exposure and outcome

Exposures were measured by assessing exposure on the risk factors including baseline characteristics, comorbidity, tobacco consumption, of the COVID-19 patients retrospectively.

Outcomes were measured by assessing the morbidity outcome in terms of cured or not-cured and the mortality outcome in terms of survivor or non-survivor.

### Ethics

The study was conducted by maintaining all kinds of ethical issues in different stages of the study. Ethical clearance was obtained from the Institutional Ethics Committee (IEC) of NIPSOM, Dhaka, Bangladesh. Informed consent was obtained from the participants by informing the purpose and procedure, expected duration, nature, and anticipated physical and psychological risks and benefits of participating. The confidentiality of data and privacy of the participants was strictly maintained. The participants were offered the right to withdraw their consent at any stage of the study. Data were stored into the computer at the central office, NIPSOM under the direct supervision of the principal investigator. Data were used anonymously for the present study only.

## Results

Out of 1187 COVID-19 patients, 1016 (85.6%) were enrolled followed by 6.6% had a wrong contact number, 3.5% didn’t attend phone calls, 3.3% were unwilling to participate and 1.0% had an incomplete interview –**Figure 1**.

**Figure 1:**
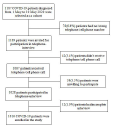
Flow chart of the study participants(COVID-19 patients)

### Baseline characteristics of COVID-19 patients

Among 1016 COVID-19 patients, the majority (64.1%) were males 89.9% were aged ≤ 59 years and the median (IQR) age was 37.0 (28–49) years. Of all, 72.6% were married and 39.6% completed their graduation, the majority (32.5%) were service holders and 18.6% were health workforce. More than two-third (69.3%) was from urban settings and around three-fourth(72.7%) was from a nuclear family. The majority (37.0%) had monthly family income between TK.10000–30000 and their average monthly family income was TK.52859(±51000.37) –**Table 1**.

**Table 1.**
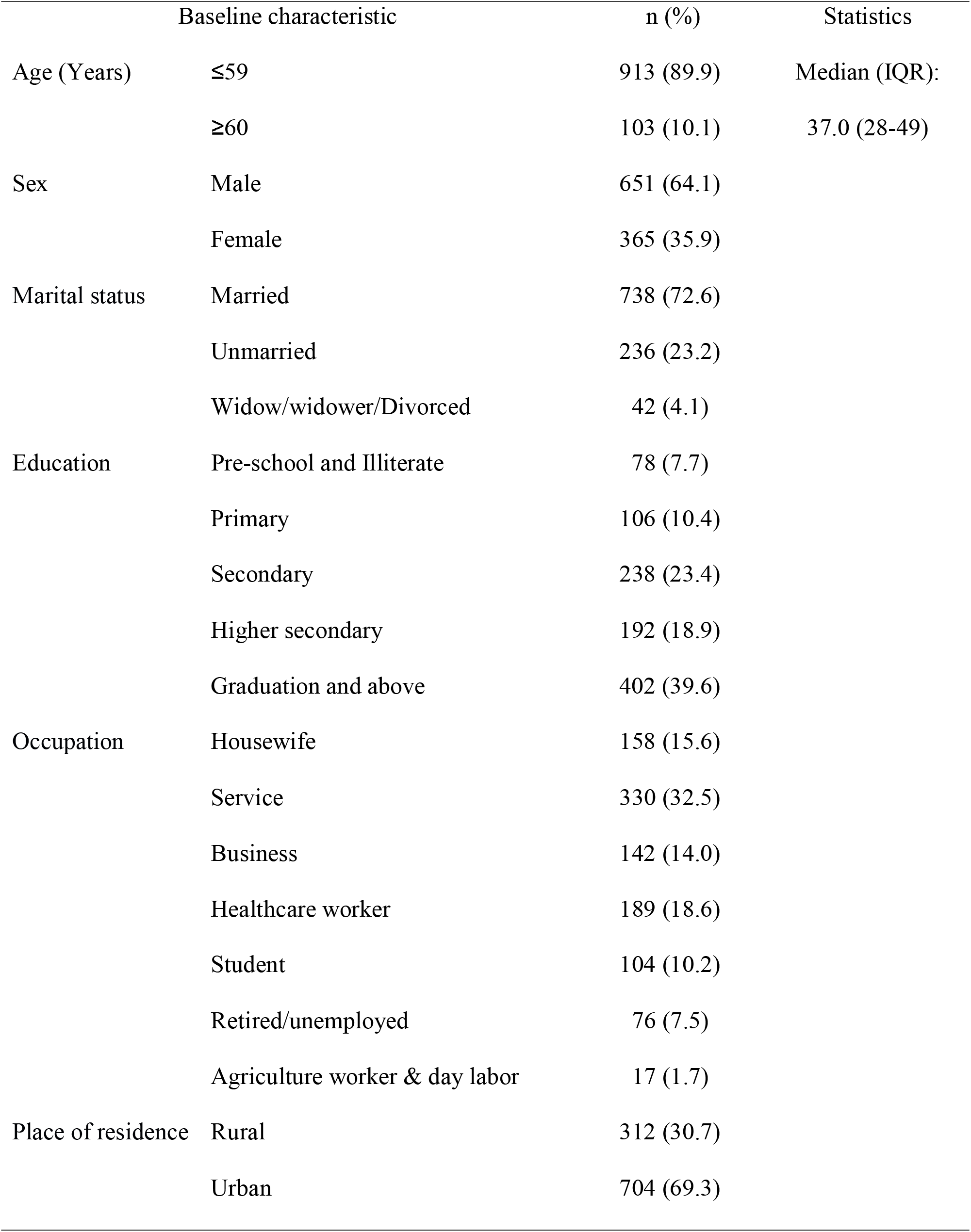
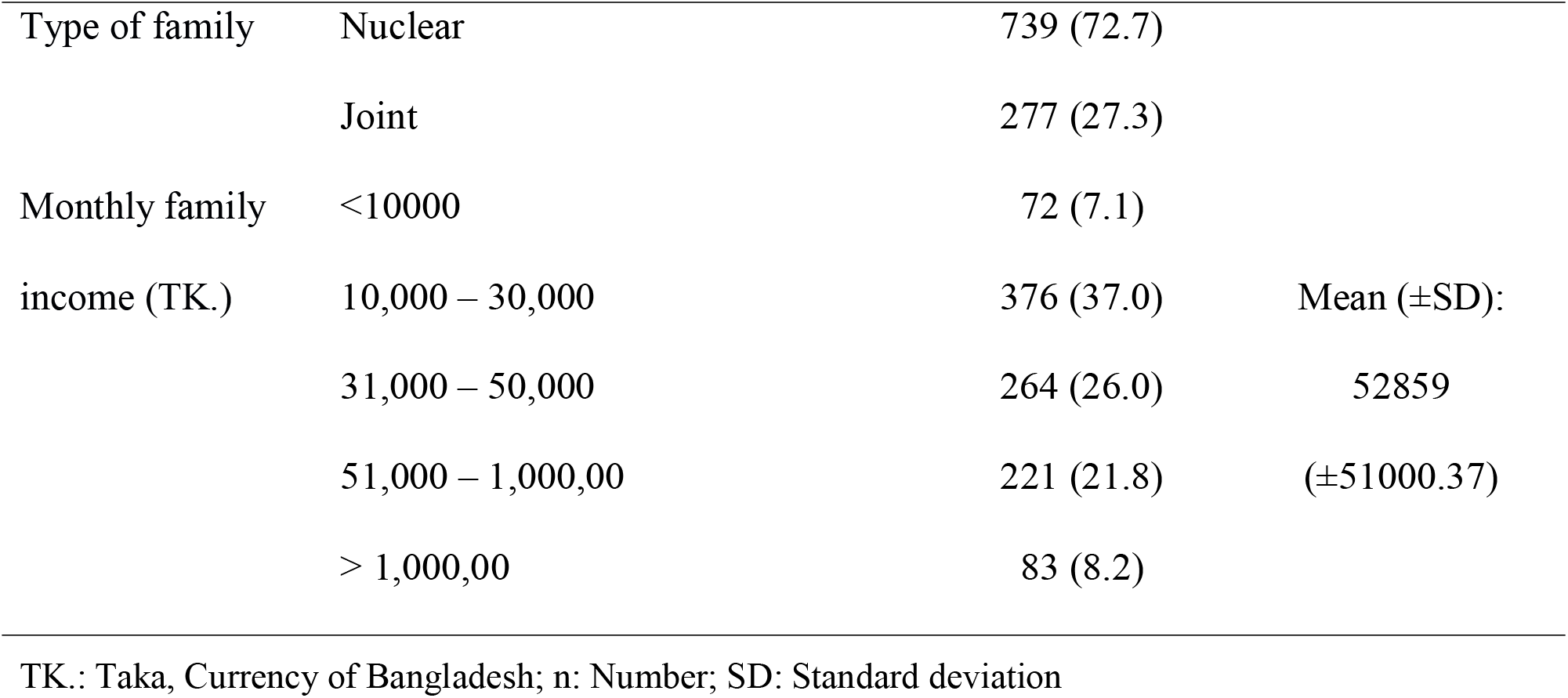
Distribution of the COVID-19 patients by baseline characteristics

|  | Baseline characteristic | n (%) | Statistics |
| --- | --- | --- | --- |
| Age (Years) | ≤59 | 913 (89.9) | Median (IQR):<br>37.0 (28-49) |
|  | ≥60 | 103 (10.1) |  |
| Sex | Male | 651 (64.1) |  |
|  | Female | 365 (35.9) |  |
| Marital status | Married | 738 (72.6) |  |
|  | Unmarried | 236 (23.2) |  |
|  | Widow/widower/Divorced | 42 (4.1) |  |
| Education | Pre-school and Illiterate | 78 (7.7) |  |
|  | Primary | 106 (10.4) |  |
|  | Secondary | 238 (23.4) |  |
|  | Higher secondary | 192 (18.9) |  |
|  | Graduation and above | 402 (39.6) |  |
| Occupation | Housewife | 158 (15.6) |  |
|  | Service | 330 (32.5) |  |
|  | Business | 142 (14.0) |  |
|  | Healthcare worker | 189 (18.6) |  |
|  | Student | 104 (10.2) |  |
|  | Retired/unemployed | 76 (7.5) |  |
|  | Agriculture worker & day labor | 17 (1.7) |  |
| Place of residence | Rural | 312 (30.7) |  |
|  | Urban | 704 (69.3) |  |
| Type of family | Nuclear | 739 (72.7) |  |
|  | Joint | 277 (27.3) |  |
| Monthly family | <10000 | 72 (7.1) |  |
| income (TK.) | 10,000 – 30,000 | 376 (37.0) | Mean (±SD): |
|  | 31,000 – 50,000 | 264 (26.0) | 52859 |
| | 51,000 – 1,000,00 | 221 (21.8) | (±51000.37) |
|  | > 1,000,00 | 83 (8.2) |  |

TK.: Taka, Currency of Bangladesh; n: Number; SD: Standard deviation

### Comorbidity

More than one third (33.9%) patients had at least 1 comorbidity. Major comorbidities were; DM (35.0%) hypertension (28.4%), COPD (16.6%), CHD (7.8%), chronic liver disease, CLD (2.5%), chronic kidney disease, CKD (4.1%), and malignant diseases (1.8%) –**Figure 2**.

**Figure 2:**
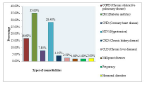
Distribution of the COVID-19 patients by types of comorbidities

### Outcomes of the COVID-19 patients

Morbidity outcomes included 35.6% cured and 64.4% not-cured on the 14^th^ day of the disease while 94.0% cured and 6.0% not-cured on the 28^th^ day. On the contrary, mortality outcomes included 97.7% survivors and 2.3% non-survivors on the 14^th^ day while 97.5% survivors and 2.5% non-survivors on the 28^th^ day –**Figure 3**.

**Figure 3:**
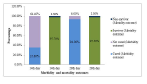
Distribution of the COVID-19 patients by outcomes(at the 14^th^ & 28^th^ day)

### Risk factors associated with outcomes of COVID-19 patients on the 14^th^ day

Regarding morbidity related risk factors, COPD was significantly (p< 0.05) higher among the not-cured (9.8%) than the cured (5.8%) patients. Regarding mortality related risk factors, elderly(60.9%) and having comorbidity (78.3%) current smokeless tobacco (SLT) use (39.1% Vs. 8.7%), COPD (13.3% Vs, 3.3%), CKD (17.4% Vs. 1.7%), and CLD (13.0% Vs. 1.1%) were significantly (p< 0.01) higher among the non-survivors than the survivors. On the other hand, DM (39.1% Vs. 16.5%), CHD (17.4% Vs. 4.0%), hypertension (34.8% Vs. 13.8%) and malignant diseases (8.7% Vs, 0.6%) were also significantly (p< 0.05) higher among the non-survivors than the survivors –**Table 2**.

**Table 2:**
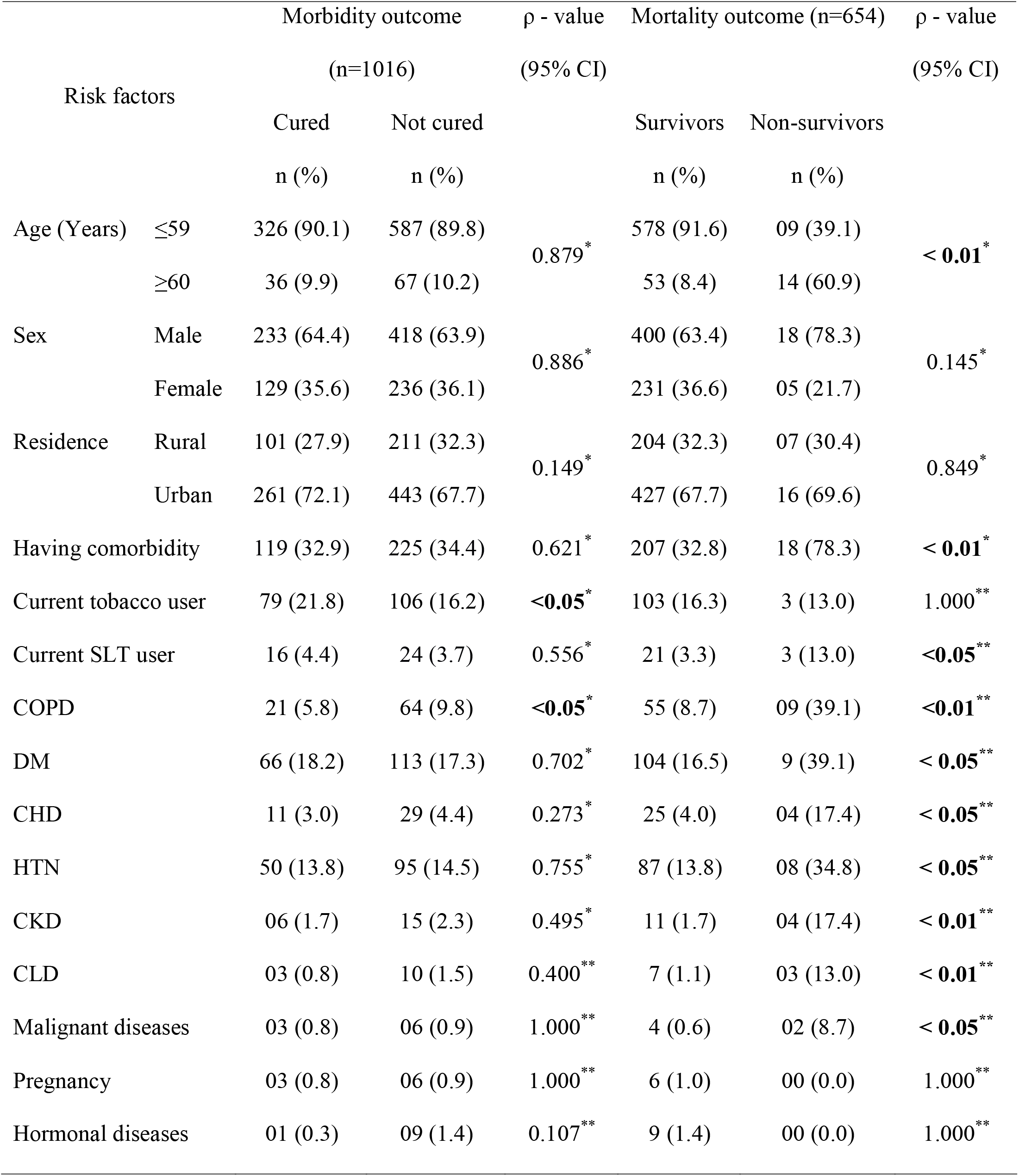
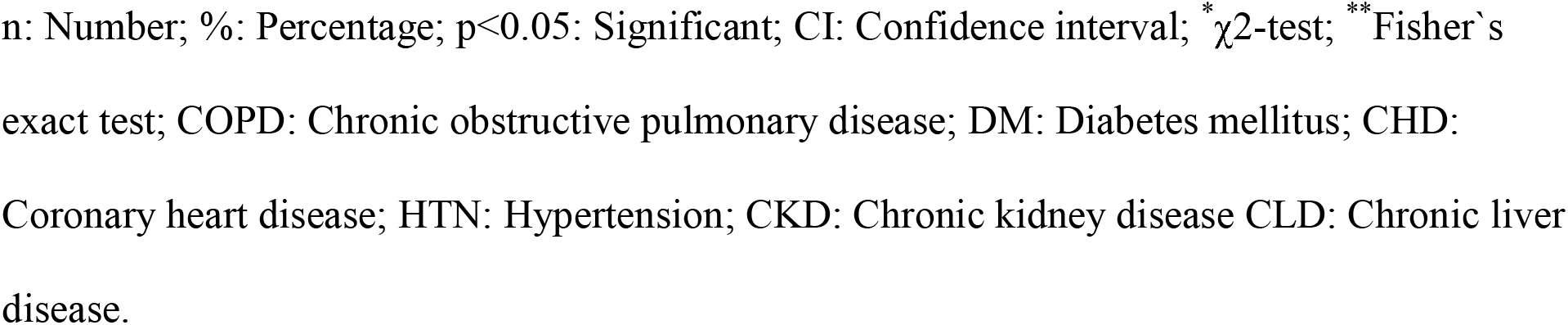
Risk factors associated with outcomes of the COVID-19 patients (On the 14^th^ day)

| Risk factors |  | Morbidity outcome |  | ρ - value<br>(95% CI) | Mortality outcome (n=654) |  | ρ - value<br>(95% CI) |
| --- | --- | --- | --- | --- | --- | --- | --- |
|  |  | (n=1016) |  |  | Survivors | Non-survivors |  |
|  |  | Cured | Not cured |  |  |  |  |
| Age (Years) | ≤59 | 326 (90.1) | 587 (89.8) | 0.879<sup>*</sup> | 578 (91.6) | 09 (39.1) | < **0.01**<sup>*</sup> |
|  | ≥60 | 36 (9.9) | 67 (10.2) |  | 53 (8.4) | 14 (60.9) |  |
| Sex | Male | 233 (64.4) | 418 (63.9) | 0.886<sup>*</sup> | 400 (63.4) | 18 (78.3) | 0.145<sup>*</sup> |
|  | Female | 129 (35.6) | 236 (36.1) |  | 231 (36.6) | 05 (21.7) |  |
| Residence | Rural | 101 (27.9) | 211 (32.3) | 0.149<sup>*</sup> | 204 (32.3) | 07 (30.4) | 0.849<sup>*</sup> |
|  | Urban | 261 (72.1) | 443 (67.7) |  | 427 (67.7) | 16 (69.6) |  |
| Having comorbidity |  | 119 (32.9) | 225 (34.4) | 0.621<sup>*</sup> | 207 (32.8) | 18 (78.3) | < **0.01**<sup>*</sup> |
| Current tobacco user |  | 79 (21.8) | 106 (16.2) | <**0.05**<sup>*</sup> | 103 (16.3) | 3 (13.0) | 1.000<sup>**</sup> |
| Current SLT user |  | 16 (4.4) | 24 (3.7) | 0.556<sup>*</sup> | 21 (3.3) | 3 (13.0) | <**0.05**<sup>**</sup> |
| COPD |  | 21 (5.8) | 64 (9.8) | <**0.05**<sup>*</sup> | 55 (8.7) | 09 (39.1) | <**0.01**<sup>**</sup> |
| DM |  | 66 (18.2) | 113 (17.3) | 0.702<sup>*</sup> | 104 (16.5) | 9 (39.1) | < **0.05**<sup>**</sup> |
| CHD |  | 11 (3.0) | 29 (4.4) | 0.273<sup>*</sup> | 25 (4.0) | 04 (17.4) | < **0.05**<sup>**</sup> |
| HTN |  | 50 (13.8) | 95 (14.5) | 0.755<sup>*</sup> | 87 (13.8) | 08 (34.8) | < **0.05**<sup>**</sup> |
| CKD |  | 06 (1.7) | 15 (2.3) | 0.495<sup>*</sup> | 11 (1.7) | 04 (17.4) | < **0.01**<sup>**</sup> |
| CLD |  | 03 (0.8) | 10 (1.5) | 0.400<sup>**</sup> | 7 (1.1) | 03 (13.0) | < **0.01**<sup>**</sup> |
| Malignant diseases |  | 03 (0.8) | 06 (0.9) | 1.000<sup>**</sup> | 4 (0.6) | 02 (8.7) | < **0.05**<sup>**</sup> |
| Pregnancy |  | 03 (0.8) | 06 (0.9) | 1.000<sup>**</sup> | 6 (1.0) | 00 (0.0) | 1.000<sup>**</sup> |
| Hormonal diseases |  | 01 (0.3) | 09 (1.4) | 0.107<sup>**</sup> | 9 (1.4) | 00 (0.0) | 1.000<sup>**</sup> |

n: Number; %: Percentage; $p < 0.05$ : Significant; CI: Confidence interval; $\chi^2$ -test; \*\* Fisher`s exact test; COPD: Chronic obstructive pulmonary disease; DM: Diabetes mellitus; CHD: Coronary heart disease; HTN: Hypertension; CKD: Chronic kidney disease CLD: Chronic liver disease.

### Risk factors associated with outcomes of COVID-19 patients on the 28^th^ day

Regarding morbidity associated risk factors, the elderly (24.6% Vs. 9.2%) were significantly (ρ < 0.01) higher among the not-cured than the cured patients. Having comorbidity (45.9% Vs. 33.1%), current SLT use (9.8% Vs. 3.6%), CKD (6.6% Vs. 1.8%) and CLD (4.9% Vs. 1.0%) were also significantly (ρ < 0.05) higher among the not-cured than the cured patients. COPD (19.7% Vs. 7.6%), CKD (6.6% Vs. 1.8%) CLD (4.9% Vs. 1.0%) were also significantly higher among the not-cured than the cured patients (< 0.05). Regarding mortality associated risk factors, the elderly (56.0% Vs. 2.8%) and having comorbidity (88.0% Vs. 22.2%) were significantly higher among the non-survivors than the survivors (11.1%), hypertension (36.0% Vs. 8.3%) and CKD (16.0% Vs. 0.0%) were also significantly higher among the non-survivors than the survivors (ρ ρ < 0.05) –**Table 3**.

**Table 3:**
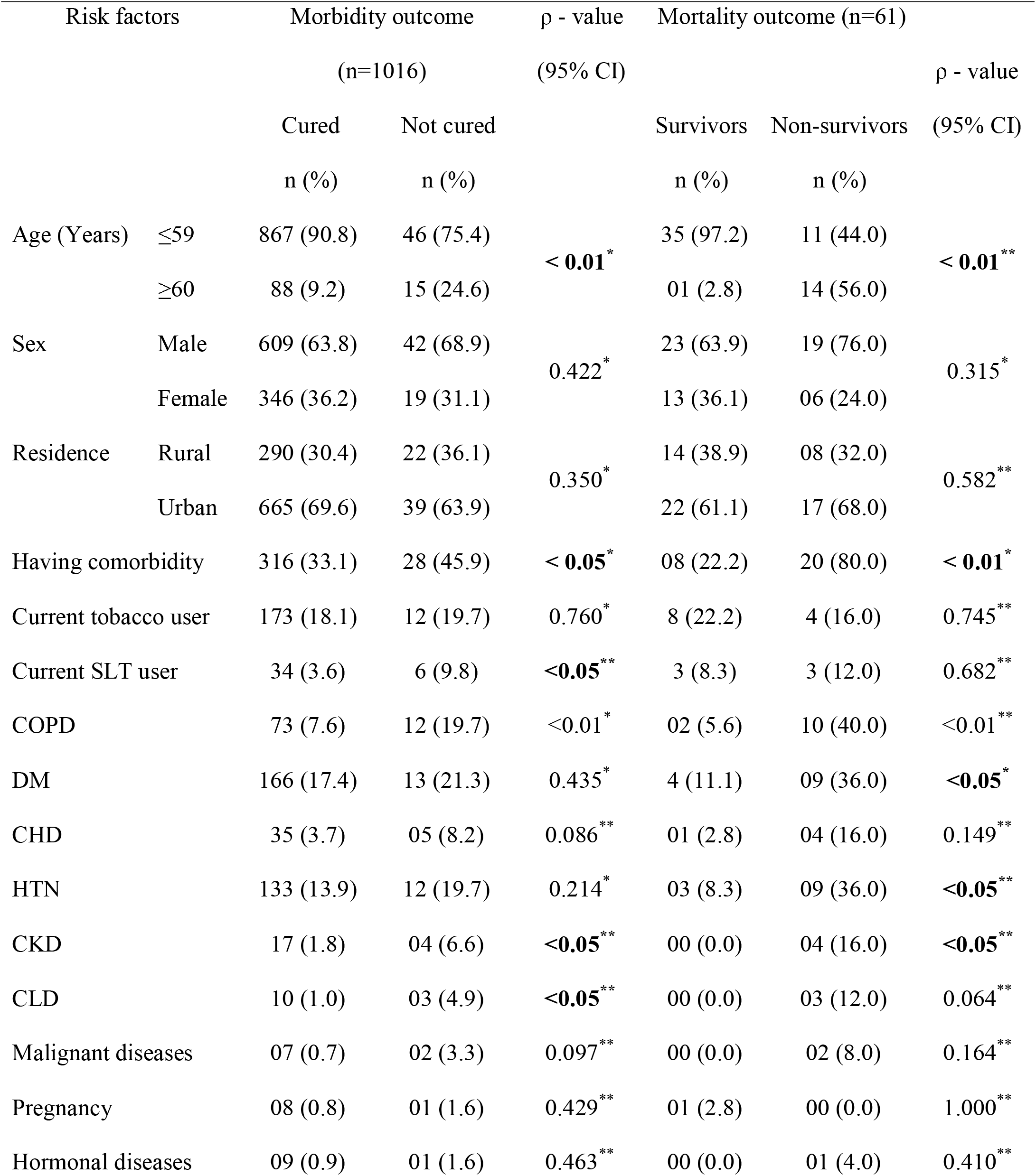
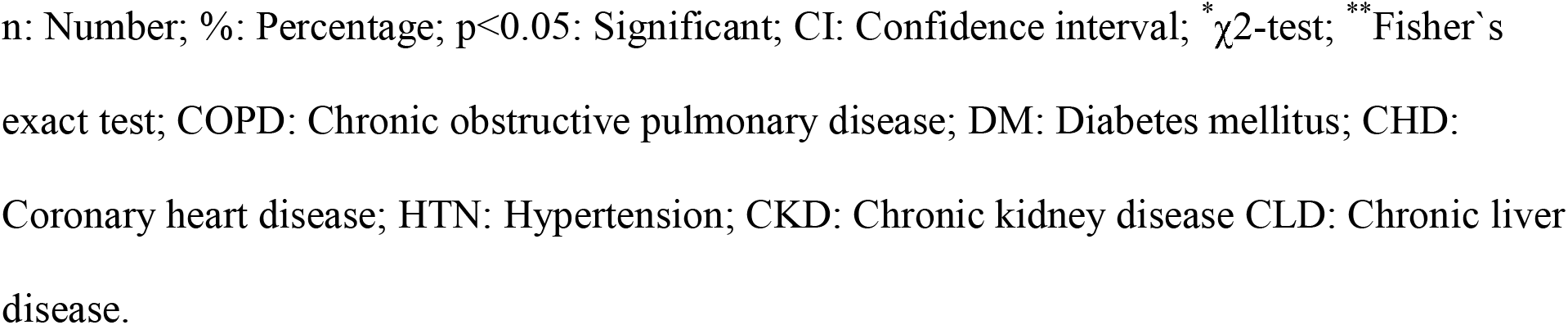
Risk factors associated with outcomes of the COVID-19 patients (On the 28^th^ day)

| Risk factors | | Morbidity outcome<br>(n=1016) | | $\rho$ - value<br>(95% CI) | Mortality outcome (n=61) | | $\rho$ - value<br>(95% CI) |
| --- | --- | --- | --- | --- | --- | --- | --- |
|  |  | Cured<br>n (%) | Not cured<br>n (%) |  | Survivors<br>n (%) | Non-survivors<br>n (%) |  |
| Age (Years) | $\leq 59$ | 867 (90.8) | 46 (75.4) | **< 0.01<sup>\*</sup>** | 35 (97.2) | 11 (44.0) | **< 0.01<sup>\*\*</sup>** |
| | $\geq 60$ | 88 (9.2) | 15 (24.6) | | 01 (2.8) | 14 (56.0) | |
| Sex | Male | 609 (63.8) | 42 (68.9) | 0.422<sup>*</sup> | 23 (63.9) | 19 (76.0) | 0.315<sup>*</sup> |
|  | Female | 346 (36.2) | 19 (31.1) |  | 13 (36.1) | 06 (24.0) |  |
| Residence | Rural | 290 (30.4) | 22 (36.1) | 0.350<sup>*</sup> | 14 (38.9) | 08 (32.0) | 0.582<sup>**</sup> |
|  | Urban | 665 (69.6) | 39 (63.9) |  | 22 (61.1) | 17 (68.0) |  |
| Having comorbidity |  | 316 (33.1) | 28 (45.9) | **< 0.05<sup>\*</sup>** | 08 (22.2) | 20 (80.0) | **< 0.01<sup>\*</sup>** |
| Current tobacco user |  | 173 (18.1) | 12 (19.7) | 0.760<sup>*</sup> | 8 (22.2) | 4 (16.0) | 0.745<sup>**</sup> |
| Current SLT user |  | 34 (3.6) | 6 (9.8) | **< 0.05<sup>\*\*</sup>** | 3 (8.3) | 3 (12.0) | 0.682<sup>**</sup> |
| COPD |  | 73 (7.6) | 12 (19.7) | **< 0.01<sup>\*</sup>** | 02 (5.6) | 10 (40.0) | **< 0.01<sup>\*\*</sup>** |
| DM |  | 166 (17.4) | 13 (21.3) | 0.435<sup>*</sup> | 4 (11.1) | 09 (36.0) | **< 0.05<sup>\*</sup>** |
| CHD |  | 35 (3.7) | 05 (8.2) | 0.086<sup>**</sup> | 01 (2.8) | 04 (16.0) | 0.149<sup>**</sup> |
| HTN |  | 133 (13.9) | 12 (19.7) | 0.214<sup>*</sup> | 03 (8.3) | 09 (36.0) | **< 0.05<sup>\*\*</sup>** |
| CKD |  | 17 (1.8) | 04 (6.6) | **< 0.05<sup>\*\*</sup>** | 00 (0.0) | 04 (16.0) | **< 0.05<sup>\*\*</sup>** |
| CLD |  | 10 (1.0) | 03 (4.9) | **< 0.05<sup>\*\*</sup>** | 00 (0.0) | 03 (12.0) | 0.064<sup>**</sup> |
| Malignant diseases |  | 07 (0.7) | 02 (3.3) | 0.097<sup>**</sup> | 00 (0.0) | 02 (8.0) | 0.164<sup>**</sup> |
| Pregnancy |  | 08 (0.8) | 01 (1.6) | 0.429<sup>**</sup> | 01 (2.8) | 00 (0.0) | 1.000<sup>**</sup> |
| Hormonal diseases |  | 09 (0.9) | 01 (1.6) | 0.463<sup>**</sup> | 00 (0.0) | 01 (4.0) | 0.410<sup>**</sup> |

n: Number; %: Percentage; $p < 0.05$ : Significant; CI: Confidence interval; $\chi^2$ -test; \*\* Fisher`s exact test; COPD: Chronic obstructive pulmonary disease; DM: Diabetes mellitus; CHD: Coronary heart disease; HTN: Hypertension; CKD: Chronic kidney disease CLD: Chronic liver disease.

Logistic regression analysis revealed that the risk of ‘not-cured’ (morbidity) was significantly higher (p< 0.05) among the elderly (AOR:2.56, 95% CI:1.31–4.99), patients with comorbidity (OR:1.72, 95% CI:1.02–2.89) and current SLT users (OR:2.95, 95% CI:1.19–7.34) on the 28^th^ day. The risk of ‘non-survivor’ (mortality) was significantly (p< 0.01) higher in the elderly (AOR: 10.14, 95% CI: 3.98–25.79) and patients having comorbidity (AOR: 4.32, 95% CI: 1.49–12.60) on the 14^th^ day. On the contrary, the risk of being non-survivor outcome was significantly higher among the elderly (AOR: 24.37, 95% CI: 2.59–229.37) and patients with comorbidity (AOR: 7.91, 95% CI: 1.91–32.70) –**Table 4**.

**Table 4:** Logistic regression analysis of the risk factors associated with morbidity (Not-cured) and mortality (non-survivor) outcomes of COVID-19 patients

| Risk factors |  | Unadjusted model |  |  | Adjusted model |  |  |
| --- | --- | --- | --- | --- | --- | --- | --- |
|  |  | OR | 95% CI | *p-value* | AOR | 95% CI | *p-value* |
| **Risk factors associated with morbidity outcome (not-cured)** |  |  |  |  |  |  |  |
| On the 28<sup>th</sup> day | Age |  |  |  |  |  |  |
|  | ≤59 | Reference |  |  | Reference |  |  |
|  | ≥60 | 3.213 | (1.72-5.99) | 0.000 | 2.559 | (1.31-4.99) | <0.01 |
|  | Having comorbidity |  |  |  |  |  |  |
|  | No | Reference |  |  | Reference |  |  |
|  | Yes | 1.716 | (1.02-2.89) | 0.042 | 1.431 | (0.83-2.47) | 0.198 |
|  | Current SLT use |  |  |  |  |  |  |
|  | No | Reference |  |  | Reference |  |  |
|  | Yes | 2.955 | (1.19-7.34) | 0.020 | 2.168 | (0.84-5.61) | 0.110 |
| **Risk factors associated with mortality outcome (non-survivor)** |  |  |  |  |  |  |  |
| On the 14<sup>th</sup> day | Age (Years) |  |  |  |  |  |  |
|  | ≤59 | Reference |  |  | Reference |  |  |
|  | ≥60 | 16.964 | (7.01-41.04) | 0.000 | 10.136 | (3.98-25.79) | <0.01 |
|  | Having comorbidity |  |  |  |  |  |  |
|  | No | Reference |  |  | Reference |  |  |
|  | Yes | 7.374 | (2.70-20.14) | 0.000 | 4.326 | (1.49-12.60) | <0.01 |
|  | Current SLT use |  |  |  |  |  |  |
|  | No | Reference |  |  | Reference |  |  |
|  | Yes | 4.357 | (1.20-15.81) | 0.025 | 2.253 | (0.53-9.61) | 0.273 |
| On the 28<sup>th</sup> day | Age (Years) |  |  |  |  |  |  |
|  | ≤59 | Reference |  |  | Reference |  |  |
|  | ≥60 | 44.454 | (5.25-378.19) | 0.001 | 24.367 | (2.59-229.37) | <0.01 |
|  | Having comorbidity |  |  |  |  |  |  |
|  | No | Reference |  |  | Reference |  |  |
|  | Yes | 14.000 | (3.99-49.16) | 0.000 | 7.908 | (1.91-32.70) | <0.01 |

CI: Confidence interval; OR: Odds Ratio; AOR: Adjusted Odds Ratio; SLT: Smokeless tobacco

### Relative Risk and Attributable Risk of outcomes of COVID-19 patients

The risk of ‘not-cured’ outcome was higher (RR:1.19, AR:11.92%) among the patients with COPD on the 14^th^ day while it was higher among the elderly (RR:2.89, AR:9.52%) and patients with comorbidity (RR:1.66, AR: 3.23%), current SLT users (RR = 2.66, AR: 9.36%), CKD (RR:3.33, AR:3.32%), CLD (RR:3.99, AR:17.29%) and COPD (RR:2.68, AR:8.85%) on the 28^th^ day. The risk of ‘non-survivor’ outcome was higher among the elderly (RR:13.63, AR:19.36%), patients with comorbidity (RR:6.86, AR: 6.83%), current SLT users (RR = 3.94, AR:9.33%), DM (RR:3.08, AR:5.38%), CHD (RR:4.54, AR:10.75%), hypertension (RR:3.14, AR:5.74%), CKD (RR:8.97, AR:23.69%), CLD (RR:9.66, AR:26.89%) and malignant diseases (RR:10.29, AR:30.09%) on the 14th day while it was also higher among the elderly (RR:3.90, AR:69.42%), patients with comorbidity (RR:4.71, AR:56.28%), DM (RR:2.08, AR:35.90%), hypertension (RR:2.30, AR:42.35%) and CKD (RR:2.71, AR:63.16) on the 28^th^ day –**Table 5**.

**Table 5:**
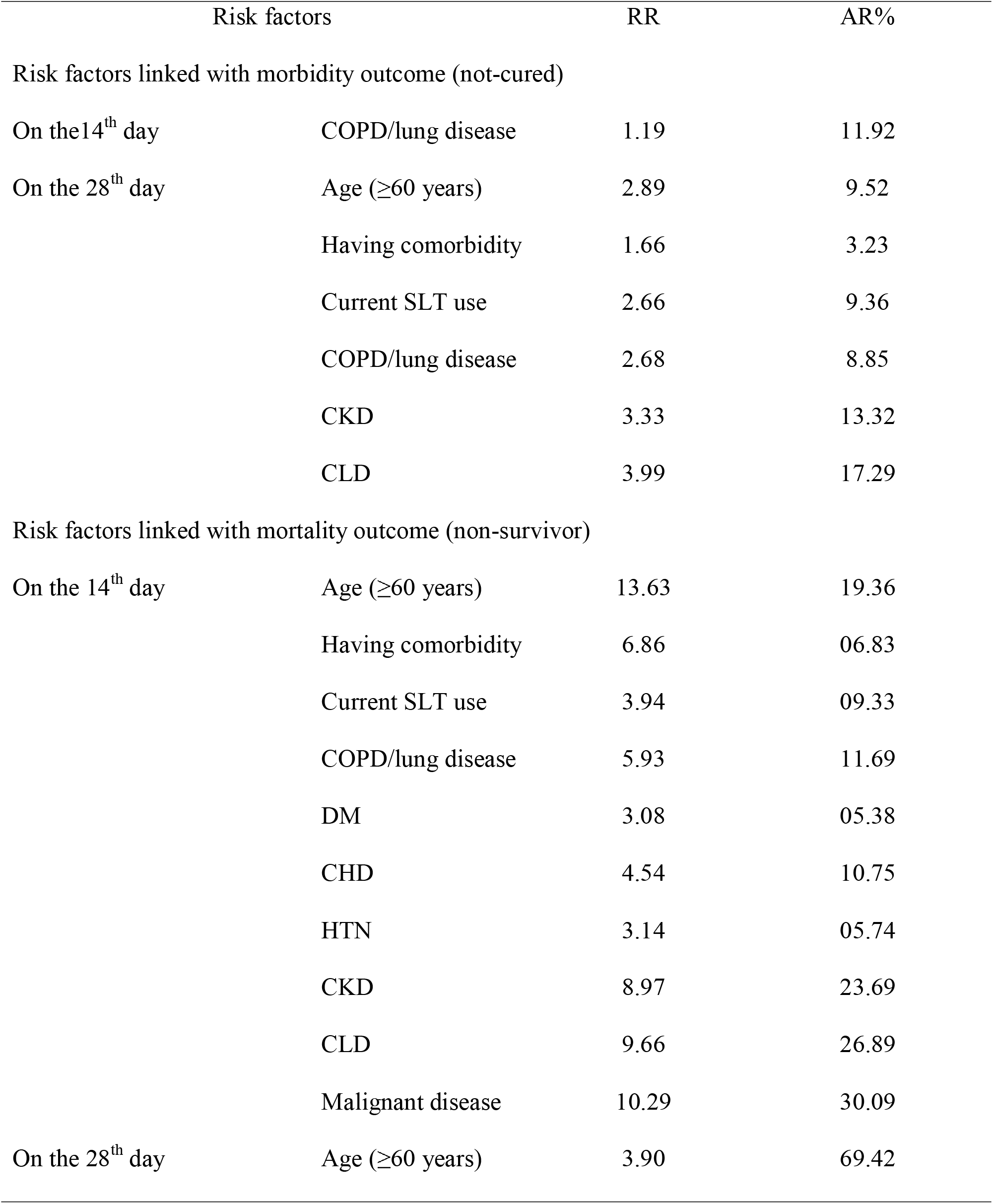
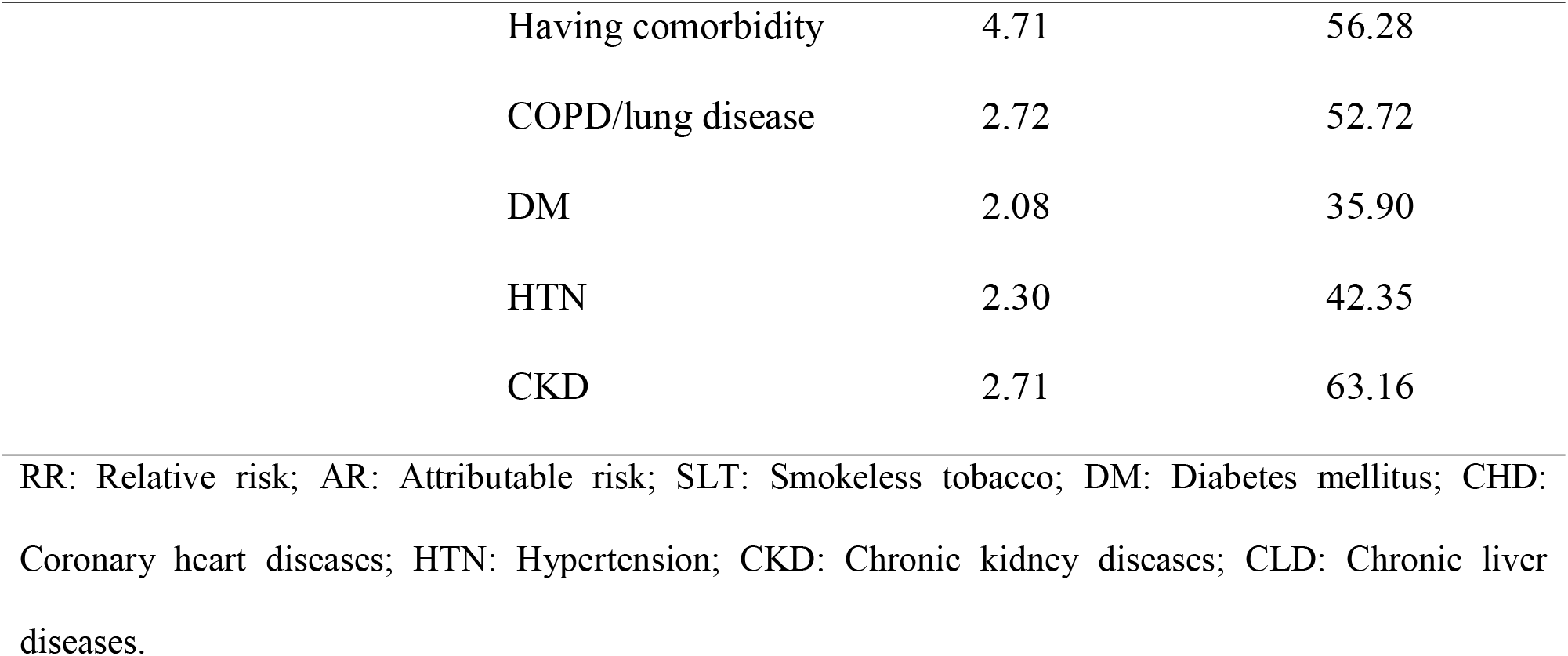
Relative Risk and Attributable Risk of morbidity (Not-cured) and mortality (non-survivor) outcomes of COVID-19 patients by selected risk factors

| Risk factors |  | RR | AR% |
| --- | --- | --- | --- |
| Risk factors linked with morbidity outcome (not-cured) |  |  |  |
| On the 14<sup>th</sup> day | COPD/lung disease | 1.19 | 11.92 |
| On the 28<sup>th</sup> day | Age ($\geq 60$ years) | 2.89 | 9.52 |
|  | Having comorbidity | 1.66 | 3.23 |
|  | Current SLT use | 2.66 | 9.36 |
|  | COPD/lung disease | 2.68 | 8.85 |
|  | CKD | 3.33 | 13.32 |
|  | CLD | 3.99 | 17.29 |
| Risk factors linked with mortality outcome (non-survivor) |  |  |  |
| On the 14<sup>th</sup> day | Age ($\geq 60$ years) | 13.63 | 19.36 |
|  | Having comorbidity | 6.86 | 06.83 |
|  | Current SLT use | 3.94 | 09.33 |
|  | COPD/lung disease | 5.93 | 11.69 |
|  | DM | 3.08 | 05.38 |
|  | CHD | 4.54 | 10.75 |
|  | HTN | 3.14 | 05.74 |
|  | CKD | 8.97 | 23.69 |
|  | CLD | 9.66 | 26.89 |
|  | Malignant disease | 10.29 | 30.09 |
| On the 28<sup>th</sup> day | Age ($\geq 60$ years) | 3.90 | 69.42 |

|  |  |  |
| --- | --- | --- |
| Having comorbidity | 4.71 | 56.28 |
| COPD/lung disease | 2.72 | 52.72 |
| DM | 2.08 | 35.90 |
| HTN | 2.30 | 42.35 |
| CKD | 2.71 | 63.16 |

RR: Relative risk; AR: Attributable risk; SLT: Smokeless tobacco; DM: Diabetes mellitus; CHD: Coronary heart diseases; HTN: Hypertension; CKD: Chronic kidney diseases; CLD: Chronic liver diseases.

## Discussion

This single centered retrospective cohort study was conducted to identify the risk factors associated with the outcomes of COVID-19 patients on the 14^th^ day and the 28^th^ day of the disease course. The study results on morbidity and mortality associated risk factors including age, comorbidity, and tobacco consumption propose decisive academic, policy, preventive, promotive, and curative strategies and interventions to combat the COVID-19 impact.

### Strength and weakness of the study

The current study is the pioneering initiative for the first time done in Bangladesh on the risk factors associated with outcomes of COVID-19 patients. This observational retrospective cohort design was scientifically apposite and feasible for identifying multiple risk factors at a single attempt. The sample size was large enough to draw a valid inference on the risk factors linked with the outcomes of COVID-19 patients considering both morbidity and mortality consequences. Though the study results might not reflect the scenario of the whole country, it offers an impression on the problem under the study with an urban-rural mix and representation of the population of the COVID-19 patients. Despite a few limitations of recall bias that emerged through a telephone-interview and wrong telephone contact number, the study findings conserve irrefutable policy implications for prevention and control of the worst outcomes of the COVID-19 patients.

### Baseline characteristics

Though the males (64.1%) have a reasonably higher risk of being affected by COVID-19 than their counterpart females (35.9%), but no significant gender difference of outcomes was revealed in our study. Similar findings were also found by other studies [7–8, 12–13] where males were being affected more than females. It is evident that the males are more involved in outdoor activities in our context and thus they are more vulnerable than the females. Though the majority(89.9%) of the patients were aged ≤ 59 years, the adverse outcomes were more prevalent among elderly patients (≥ 60 years). Urban prevalence (69.3%) was higher than the rural; it could be argued that unplanned urbanization, higher population density, and industrialization in the urban areas increase disease transmission and prevalence. Moreover, more aware urban people undergo laboratory tests for COVID-19 more than the rural people. By occupation, lion shareholders were service holders (32.5%) and health workforce (18.6%). It is evident that the service holders including bankers, security forces, police, and community forces provide various emergency services and are exposed to COVID-19 infection. The health workforce including doctors, nurses, and supporting staff are more vulnerable for the COVID-19 as they have to provide healthcare in direct contact with the patients. Besides, poor quality and quantity of personal protective equipment (PPE) also aggravate their vulnerability for the disease.

### Comorbidity and outcomes of COVID-19 patients

Of all, 33.9% patients had diverse comorbidities including DM (35.0%), hypertension (28.4%), COPD (16.6%), and CHD (7.8%). Another study conducted in Wuhan, China [8] also identified DM, hypertension, COPD, and CHD as major comorbidities with the COVID-19 patients. A large difference (64.4% Vs. 6.0%) of morbidity outcome (not-cured) while a small difference(2.3% and 2.5%) of mortality outcome was revealed between the 14^th^ and 28^th^ day of the disease course respectively. It reflects that the worst mortality outcome occurs within the first 14 days while most of the patients come round by the second 14 days of the disease. So, patient management for the first 14 days must be emphasized and prioritized considering associated risk factors to reduce the morbidity and mortality burden of COVID-19. A study conducted in China revealed different findings, where the death rate was 3.6% among Chinese patients and 1.5% among patients outside China [14]. This discrepancy demands comprehensive comparative studies, but it may be the result of differences in host factors like body immunity, comorbidity, food habit, etc. and agent factors like genomes, virulence, transmission probability, etc. between the geographies.

### Risk factors associated with morbidity outcome

COPD (RR = 1.19, AR% = 11.92) and elderly (AOR = 2.56, RR = 2.89) were significantly associated with the ‘not-cured’ morbidity outcome on the 14^th^ and 28^th^ day of the disease. Other relevant studies [12, 15] showed similar findings in respect of age, COPD, and morbidity outcomes. It is established that poor body immunity of the elderly patients instigates the worse progression and adverse outcomes of the COVID-19. Current SLT use (OR = 2.96, RR = 2.66), CKD (RR = 3.33), and CLD (RR = 3.99) were found independently associated with morbidity (‘not-cured’) outcome of COVID-19 on the 28^th^ day. It could be explained by the fact that comorbidity increases the severity of COVID-19 and prolong the morbid condition.

### Risk factors associated with mortality outcome

Elderly (AOR = 10.14, RR = 13.63) and having comorbidity (AOR = 4.33, RR = 6.86) were significantly associated with the mortality outcome of COVID-19 on the 14^th^ day of the disease. A study conducted in Wuhan, China [8] also showed similar findings. Though the majority of the patients of our study were in the middle age group, the mortality rate was higher among the elderly. It can be justified by the fact that compromised body immunity of the elderly patients having comorbidity could not win the fight with COVID-19 rather confront the worst outcome. Mortality outcome (non-survivor/death) was also significantly associated with COPD (RR = 5.93), DM (RR = 3.08), CHD (RR = 4.54), hypertension (RR = 3.14), CKD (RR = 8.97), CLD (RR = 9.66), and malignant diseases (RR = 10.29) on the 14^th^ day. It is recognized that all the comorbidities weaken the immune system of the patients and aggravate the worst mortality outcome. Though current smoking didn’t show any significant association with the morbidity and morbidity outcomes and current SLT use was independently associated with the mortality on the 14^th^ day of the disease (OR = 4.36, RR = 3.94). It is reported that 27.5% adult people (26.9 % men, 28.1% women) of Bangladesh take ‘zarda’, ‘khoinee’, ‘gul’, ‘sadapata’, ‘nossi’, etc., which are local smokeless tobacco products [16]. It is argued that smokeless tobacco induces pathophysiological changes in the upper respiratory tract, which makes the virus more progressive and aggressive to aggravate acute respiratory distress syndrome and mortality. Further intensive research and analysis could be conducted to establish the scientific arguments on this association in the Indian sub-continent including Bangladesh.

On the 28^th^ day, mortality outcome was significantly associated with the elderly (AOR = 24.367, RR = 3.90) and having comorbidity (AOR = 7.908, RR = 4.71). Another retrospective study conducted in Wuhan; China [8] also revealed that higher median age (69.0 years) of the non-survivors than the survivors (52.0 years). The study also found comorbidity more in the non-survivors (67%) than the survivors (40%). Mortality outcome was also significantly associated with COPD (RR = 2.72), DM (RR = 2.08), hypertension (RR = 2.30), and CKD (RR = 2.71). These findings were consistent with other relevant studies conducted in different countries [8,12,14]. It is claimed that the comorbidity deteriorates the defensive mechanism of the patients and worsen the mortality outcome of COVID-19 patients.

### Policy and public health implications

The identified risk factors associated with the outcomes of the COVID-19 patients conserve crucial policy implications for prevention and control of the morbidity and mortality burden of the disease. The study results could contribute to strengthen and reorganize the health care delivery system of the country for providing need-oriented and prioritized services to COVID-19 patients emphasizing the first fourteen days of the disease. The study findings could also contribute to devising effective strategies for the provision of comprehensive health care to COVID-19 patients with comorbidity. Policymakers, health care managers, and relevant stakeholders may use the study findings to revise the national treatment guidelines considering the risk factors, adverse outcomes, and disease course of the COVID-19.

## Data Availability

The data for the study is available by contacting the corresponding author upon request.

## Acknowledgments

The authors are indebted to all the staff of the central laboratory of NIPSOM for their technical assistance in data generation. We are also obliged to all the COVID-19 patients and their families for participating in the study.

## Data availability statement

The data for the study is available by contacting the corresponding author upon request.

